# A cross-sectional study on hypertension and diabetes care cascade and prevalence of noncommunicable diseases (NCD) risk factors in Tamil Nadu: rationale and methods

**DOI:** 10.1101/2025.03.24.25324492

**Authors:** Archana Ramalingam, Joshua Chadwick, Manjula Neelavanan, G Anusuya, R Devendhiran, Mosoniro Krina, Vettrichelvan Venkatasamy, Emily Devasgayam, Surya Joseph, Sabarinathan Ramaswamy, Elavarasu Govindasamy, Kalaimani, Ashok Kumar, D Chokkalingam, Dinesh Durairajan, Divya Jilumudi, V Harshavardhini, Vidya Viswanathan, K Krishnaraj, Jerard Maria Selvam, T S Selvavinayagam, Arun Thamburaj, S Vineeth, P Senthil Kumar, Prabhdeep Kaur

## Abstract

**Background:** Noncommunicable diseases (NCDs) account for over 65% of all deaths in India, as per the Global Burden of Disease Report, 2019. In Tamil Nadu, a southern state in India, more than 75% of all deaths were due to NCDs. To counter the problem, the government of Tamil Nadu launched a flagship project called Makkalai Thedi Maruthuvam (MTM), a scheme that provides a comprehensive range of home-based services and doorstep delivery of drugs for NCDs. We propose to undertake a representative survey suitably designed using the WHO STEP-wise approach to estimate the proportion of individuals with hypertension or diabetes with controlled blood pressure or blood sugar in Tamil Nadu, 2024.

**Objectives:** The primary objective is to estimate the proportion of of individuals with hypertension or diabetes with controlled blood pressure or blood sugar control among individuals aged 18-69 years. The secondary objective is to estimate the prevalence of behavioural and biological risk factors for NCDs.

**Methods and analysis:** This cross-sectional study will be conducted in all districts of Tamil Nadu. We will collect data on socioeconomic status, behavioural risk factors, and physical measurements. Data will be collected on the history of screening, diagnosis, and treatment services for diabetes, hypertension, and cancer screening for oral, breast, and cervix adopted from the WHO STEPS questionnaire. Key indicators related to the burden and continuum of care for hypertension and diabetes will be estimated using complex sample wieghted analysis to estimate percentages with 95% confidence intervals.

**Ethics and dissemination:** This study proposal was approved by the institutional ethics committee. The results of our study will be shared through the manuscripts in peer-reviewed journals. Additionally, the findings will be presented to the relevant public health authorities in India for drafting NCD policies and management.

## Introduction

Noncommunicable diseases (NCDs) account for over 65% of all deaths in India, as per the Global Burden of Disease Report, 2019.[1] Though the epidemiological transition to a predominance of NCDs happened as early as 2003, it has not been uniform for all states of India.[2] Most NCDs are attributed to behavioural risk factors like unhealthy diet, physical inactivity, tobacco and alcohol use, and biological risk factors like high blood glucose, high blood pressure, high cholesterol, and high body mass index. Estimating the burden of these risk factors through the World Health Organization’s (WHO) standard STEPS survey will guide public health action for preventing NCDs.[3]

In 2019, in Tamil Nadu, more than 75% of all deaths were due to NCDs and high blood pressure alone led to nearly 22% of all deaths.[1] Since 2014, to counter the growing NCD epidemic, the Tamil Nadu government has adopted several multi-pronged hypertension strategies at the facility level, achieving a substantial improvement. The World Bank funded “Tamil Nadu Health System[4] Reform Program (TNHSRP) was initiated in 2019. TNHSRP aims to strengthen the institutional mechanisms and improve the management of NCDs. Improvement in population-level glycaemic control and blood pressure control were the key targets of TNHSRP. To serve as a baseline for TNHSRP, a representative survey of the prevalence of NCD risk factors in Tamil Nadu was done using the WHO-STEPS (stepwise approach to surveillance) survey in 2020.[5] The survey found that 40% of the men were current smokers, 19.3% of all individuals currently used alcohol, 15.7% were physically inactive, and 83% consumed inadequate amounts of fruits and vegetables in their diet. Raised blood pressure was seen in 31.4% of individuals, and nearly 40% had a high body mass index. Of all the individuals with hypertension, only 7% had their blood pressure under control. Glycaemic control was seen in 10.8% of all individuals with diabetes.

Immediately after the survey, the COVID-19 pandemic struck the world. The COVID-19 pandemic brought about newer challenges in ensuring the continuum of care for NCDs. There are also several studies that have documented that COVID-19 has led to an increase in the incidence of NCD risk factors like obesity and diabetes.[6–8]

To counter the problem, the government of Tamil Nadu launched a flagship project called Makkalai Thedi Maruthuvam (MTM), which provides a comprehensive range of home-based services and doorstep delivery of drugs for NCDs.[9] Having understood that periodic monitoring of the trends of the risk factors for NCDs is vital for understanding the impact of interventions done for the prevention and control of NCDs, the government of Tamil Nadu, has commissioned a second round of NCD STEPS survey. This will be critical to the future planning of strategies for NCDs in the state. With this background, we propose to undertake a representative survey suitably designed using the WHO STEP-wise approach to estimate the proportion of individuals controlled blood pressure and blood sugar in Tamil Nadu. This study will also estimate the prevalence of behavioural and biological risk factors for NCDs.

## Methods and analysis

### Objectives

Primary objective

Among individuals aged 18-69 years in Tamil Nadu,

- To estimate the proportion of individuals with controlled blood pressure among those with hypertension
- To estimate the proportion of individuals with controlled blood sugar among those with diabetes.

Secondary objective

- To estimate the prevalence of behavioural and biological risk factors for NCDs among individuals aged 18-69 years.

### Study design and setting

This will be a representative cross-sectional study done among individuals aged 18-69 years in the state of Tamil Nadu. Tamil Nadu is the southernmost state of India with an estimated population of more than 80 million.[10] Nearly half of the population lives in urban areas, as per the 2011 census.[11]

### Study Population

#### Inclusion criteria

- Adults aged 18-69 years.
- Staying for more than six months in the selected household.

#### Exclusion criteria

- Pregnant women and post-partum women up to 42 days post-delivery.

### Sample size calculation

**Table 1:**
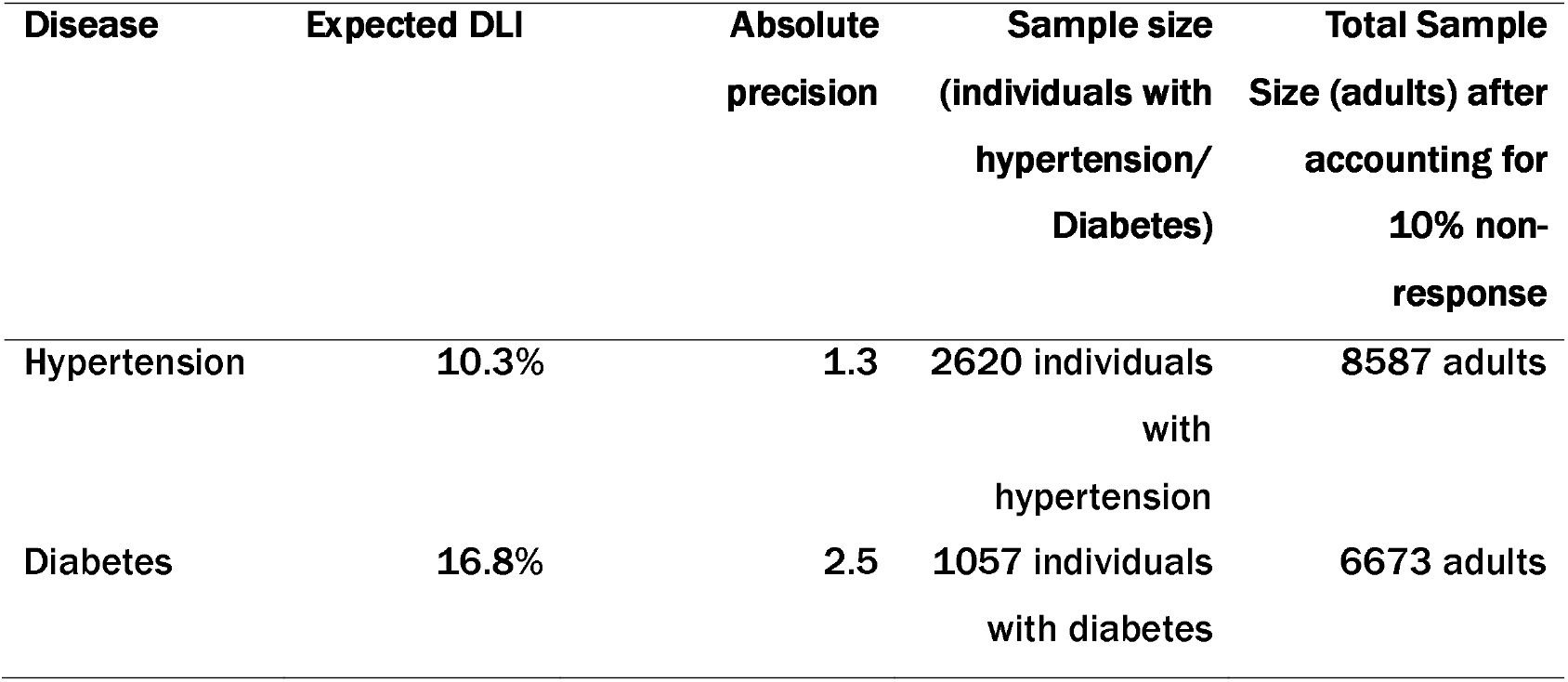
Sample size calculation for the Tamil Nadu STEPS survey, 2023-24

| Disease | Expected DLI | Absolute precision | Sample size (individuals with hypertension/ Diabetes) | Total Sample Size (adults) after accounting for 10% non-response |
| --- | --- | --- | --- | --- |
| Hypertension | 10.3% | 1.3 | 2620 individuals with hypertension | 8587 adults |
| Diabetes | 16.8% | 2.5 | 1057 individuals with diabetes | 6673 adults |

The expected proportion of individuals with controlled blood pressure among those with hypertension in Tamil Nadu as per the targets of TNHSRP was 10.3%. Therefore, to estimate the proportion of individuals with controlled blood pressure at 10.3% with an absolute precision of 1.3, 95% confidence level, and a design effect of 1.25 (multistage sampling), the minimum number of individuals with hypertension that must be surveyed was estimated to be 2620 (Open Epi).[12] Considering the prevalence of hypertension to be 33.9%,[5] the minimum number of adults to be surveyed is 7729 adults. After adjusting for 10% non-response, the total number of adults to be surveyed was 8587 adults. We rounded off the sample size to 8880 and plan to cover the same by surveying 60 adults each from 148 clusters. The calculated sample size will be sufficient to estimate the expected proportion of adults with blood sugar control (16.8%) among those with diabetes in Tamil Nadu.

### Sampling strategy

Multi-stage cluster sampling method will be adopted **Figure 1**].

**Figure 1.**
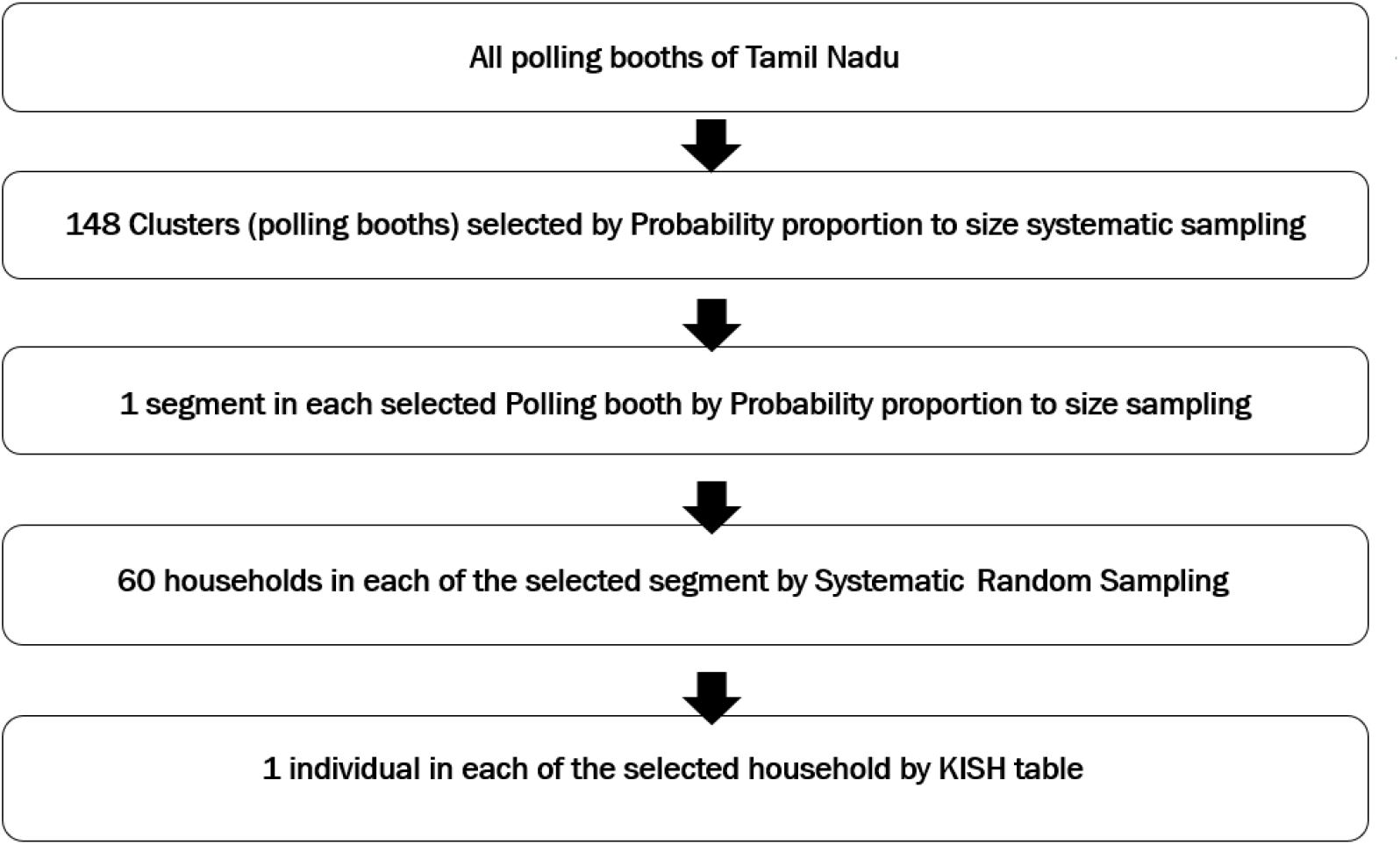
Sampling design for STEPS survey, 2024.

We will use the publicly available list of electoral booths in Tamil Nadu, 2023 (Quarter 2) as the sampling frame and choose electoral booths as clusters.[13] As per the list, there are a total of 68,036 electoral booths in Tamil Nadu and 6,23,33,925 (86.3%) adults registered as voters. [14]

#### Stage 1

Based on the total number of voters in the selected electoral list and the average size of each household in a given district (based on previous census data), we will calculate the number of households in each electoral booth. There are a few electoral booths with less than 120 households. Without disturbing the sequence or order of the electoral booth as mentioned in the publicly available list (in terms of assembly constituency name & number, and part number), we will merge electoral booths with less than 120 households with the adjacent electoral booth. Thus, we will ensure a minimum of 120 households in each electoral booth to meet the selection of a minimum of 60 households in each electoral booth.

After merging, we will use the final merged electoral booth list without disturbing the order as the sampling frame. We will select 148 electoral booths using probability proportion to size (number of households as size) systematic sampling (PPSS).

#### Stage 2

After the selection of 148 clusters (electoral booth), we plan to segment each cluster into 1 to 4 segments depending on the total number of households. After segmenting, we will select 1 segment in each electoral booth using probability proportion to size (number of households as size) sampling (PPS), depicted in **figure 2**.

**Figure 2.**
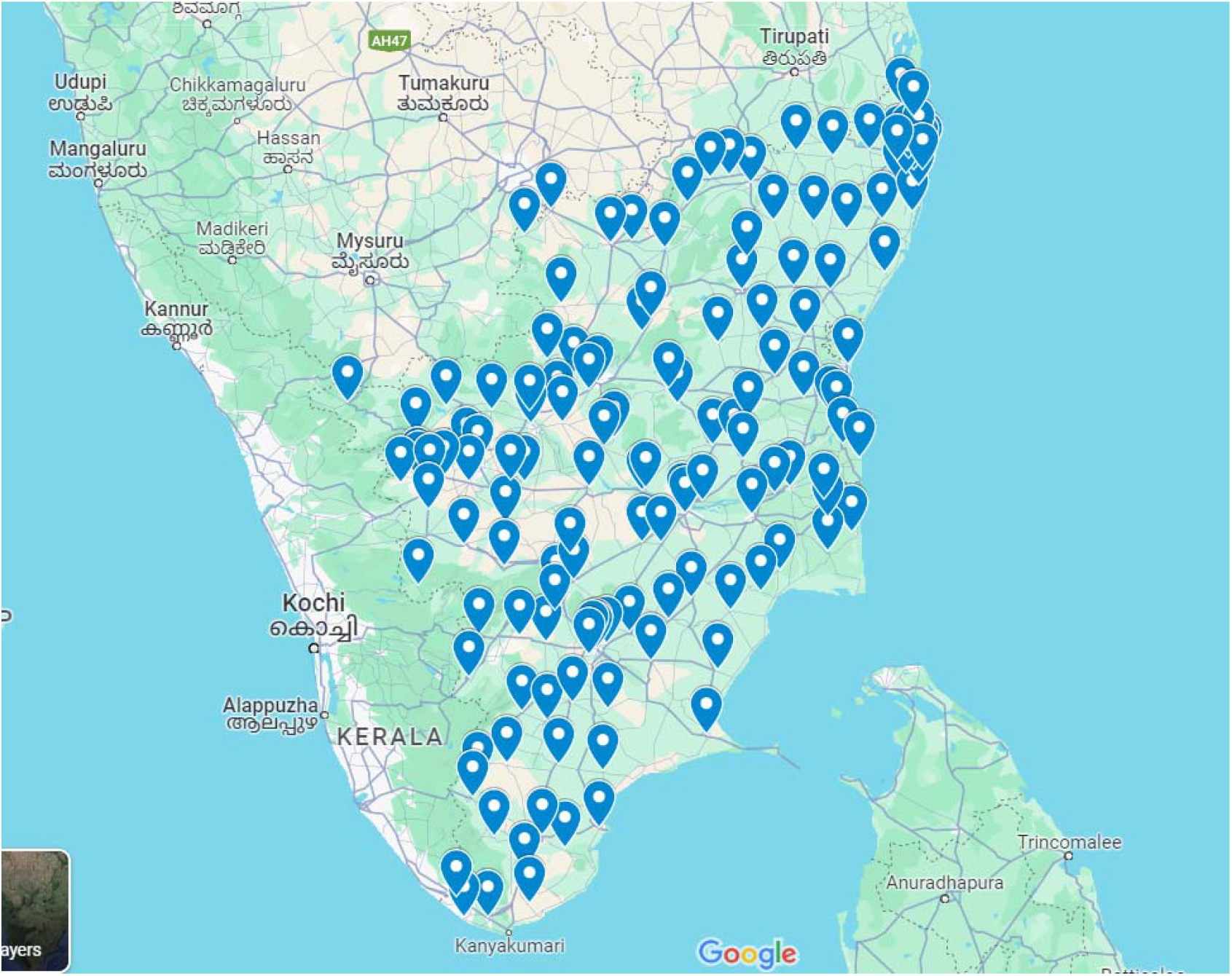
Geographical distribution of sampled clusters for TN STEPS Survey, 2023-24.

#### Stage 3

After the selection of 1 segment from each of the selected clusters, we will list all the households in the selected segment. To begin the household listing, we will start from the first household of the selected segment, assign a unique number to each household and will always keep moving in the left direction till the listing is completed. During the household listing, we will enquire about the presence of at least one adult aged 18–69 years. The households that have eligible participants for the survey will be marked and considered for selection in the third stage. The household data for listing will be collected electronically using tablets on an ODK form. The list of all the households of the selected segment will be uploaded by the field team to the ICMR-NIE server.

After the household list is uploaded to the server, 60 eligible households from each segment will be selected using systematic random sampling with the help of an in-built SYS software in the ICMR-NIE portal.

#### Stage 4

Once the 60 households with eligible participants are selected in each segment by the software, they will not be changed at any time. The list of selected households will be downloaded by the central statistical team at ICMR-NIE and the same will be shared with the field data collection teams.

After the list of 60 households is downloaded and taken as a printout, the data collection team will visit all the selected 60 households. In each household, the field team will enumerate all the eligible adults aged 18–69 years and will enter the data in the ODK form. The ODK form will have an in-built KISH algorithm which will select one individual per household for the study.

In each of the first 3 stages of the sampling, a decimal random number will be used and will be fixed for reproducibility.

### Data collection

The WHO STEPS Survey has 3 steps:

STEP I: Questionnaire-based data collection.

STEP II: Anthropometry and blood pressure measurement.

STEP III: Biochemical measurements.

We will deploy 14 data collection teams, each with a supervisor and four data collectors, to cover a cluster in five days. We will train the data collection team for a week using lectures, role-play, hands-on practical sessions, and group discussions. After the training, each team will collect data for the first cluster under supervision by the core team at ICMR-NIE. Interviews will be conducted in person, with data being gathered using handheld devices through ODK software.

We will use the adapted WHO STEPS questionnaire for data collection. Data on socioeconomic status, behavioural risk factors, history of screening, diagnosis, and treatment for diabetes and hypertension will be collected. Detailed information for each question is available in **Annex 1**.

### Tools

To assess physical measurements such as height, weight, and waist circumference (WC), standardized equipment will be utilized—namely, a portable stadiometer (SECA 213), a digital scale (SECA 803), and a measuring tape (SECA, 201, Seca GmbH Co, Hamburg, Germany), following WHO-STEPS guidelines. Participants who give consent will have their height and weight measured while they are barefoot and wearing light clothing, standing upright with their heads straight and arms at their sides. The body mass index will be calculated by dividing the weight in kilograms by the square of the height in meters. The WC will be measured at the midpoint between the lowest rib and the top of the hip bone, at the end of a normal exhalation, with the participant’s arms relaxed at their sides, and the measurement will be recorded to the nearest 0.1cm. Blood pressure will be measured using a standardized automatic machine (OMRON HEM–1320, Omron Corporation, Kyoto, Japan). Three readings will be taken while the participant is seated, with a five-minute rest between each reading, and the average of the last two readings will be considered.

The selected participant will be advised to be fasting the subsequent morning. Fasting blood sugar will be measured using a glucometer-based (*Accu-Chek Instant S blood glucose meter, Roche Diagnostics, Germany*) capillary fasting blood glucose (dry strip method) testing method. All instruments will be regularly calibrated, and the survey teams will maintain calibration logbooks.

When a household is found to be locked, or the selected respondent is not available during the initial visit, a notice regarding the visit will be left behind. At least three additional visits will be scheduled, accommodating the participants’ available times. If at the end of three visits, the participant is not available, then it will be considered as non-response. Cases of refusal or partial completion will also constitute non-response.

**Table 2:** Operational Definitions[15–17]:

| **Outcomes** | **Hypertension** | **Diabetes** |
| --- | --- | --- |
| **Definition** | Anyone with AT LEAST ONE of the following:<br>i.Average of second and third BP reading: Systolic Blood Pressure (SBP) ≥ 140 OR Diastolic Blood Pressure (DBP) ≥ 90 mmHg<br>ii. Currently taking anti-hypertensive medication | Anyone with AT LEAST ONE of the following:<br>i.Fasting capillary Glucose ≥ 126 mg/dl<br>ii.Currently taking anti-diabetic medication (either oral drugs or insulin) |
| **Awareness** | An individual with hypertension who reports AT LEAST ONE of the following:<br>a) Self-reported history of previously being diagnosed with hypertension by a health professional.<br>b) Currently taking anti-hypertensive medication | An individual with Diabetes Mellitus who reports AT LEAST ONE of the following:<br>i.Self-reported history of previously being diagnosed with Diabetes Mellitus by a health professional<br>ii. Currently taking anti-diabetic medication (either oral drugs or insulin) |
| **Treatment** | An individual with hypertension who is taking anti-hypertensive medication for at least two weeks preceding the interview | An individual with diabetes who is taking anti-diabetic medication (either oral drugs or insulin) for at least two weeks preceding the interview |
| **Control** | SBP <140 AND DBP <90 mmHg in an individual with hypertension | Fasting capillary Glucose < 126 mg/dl in an individual with Diabetes mellitus |

Also, the study sampling methods are based on the publicly available polling booth data which has updated information about the target adults for STEPS surveys. This is crucial in the context that the last census was done in 2011 in India.

Further, this study will be critical to plan future strategies for NCDs in Tamil Nadu and this is particularly important given the significant differences among states, allowing for tailored adjustments to the NCD action plan.[18,19]

### Limitations

We could not include serum cholesterol and HbA1c measurements due to logistic limitations, but studies have shown that a single blood glucose measure may be a reasonable alternative to HbA1c for community surveys.[20]

The study is dealing with health behaviours, and hence, social desirability bias may affect study results. Also, since the study is a cross-sectional one, it will help only in hypothesis generation about factors related to the stages of the care cascade.

### Outcome

This study is anticipated to significantly contribute to our understanding of the burden of NCD in Tamil Nadu by estimating the burden of NCD risk factors. Furthermore, it aims to shed light on the health-seeking behaviour of individuals diagnosed with diabetes and hypertension. Critically, it will assess the early impact of interventions such as the MTM scheme, focusing on their effectiveness in bridging the screening and treatment gaps for individuals suffering from diabetes and hypertension. Through this comprehensive approach, the study seeks to inform and enhance health policy and practice related to NCD management and prevention in Tamil Nadu and other countries with similar settings.

### Ethics and dissemination

The final study protocol, including the final version of the other essential documents, has been approved in writing by the Institutional Human Ethics Committees of the government of Tamil Nadu and ICMR-NIE. All participants will be provided information regarding the survey using a participant information sheet, and informed consent will be obtained. Confidentiality of the participants’ identity will be maintained by assigning a unique ID. Only summary statistics will be disseminated for scientific and administrative purposes. All the study procedures will be done free of cost at the participants’ doorstep. Those detected to have Diabetes Mellitus and hypertension will be referred to the nearest government health facility for further management. The results of our study will be shared through the manuscripts in peer-reviewed journals. Additionally, the findings will be presented to the relevant public health authorities in India for drafting NCD policies.

## Data Availability

All data produced in the present study are available upon reasonable request to the authors

## Acknowledgements

We thank the Directorate of Public Health and Preventive Medicine, Government of Tamil Nadu for their collaboration. We are also grateful to our colleague Mr P Kamaraj (PrinicpalTechnical Officer, ICMR-NIE) for his insightful discussions and valuable feedback about sampling, sample size and complex sample weighted analysis. His contributions greatly enhanced the quality of our work. We would like to acknowledge the STEPS (round 2) field supervisors for deligently selecting the segment from the publically available electoral booth.

## Credit Authorship contribution statement

Archana Ramalingam, Joshua Chadwick: Conception of the work, data analysis and interpretation, critical revisew and final approval of the version. Manjula Neelavanan, Anusuya Anu, Devendhiran R, Mosoniro Krina : Data collection, analysis and interpretation, final approval of the version. Vettrichelvan Venkatasamy: Data collection and interpretation, final approval of the version. Emily Devasgayam: Data collection and interpretation, final approval of the version. Surya Joseph: Data collection and interpretation, final approval of the version. Sabarinathan Ramaswamy: Data collection, analysis and interpretation, final approval of the version. Elavarasu Govindasamy: Data analysis and interpretation, final approval of the version. Kalaimani I: Data analysis and interpretation, final approval of the version. Ashok Kumar: Data analysis and interpretation, final approval of the version. Chokkalingam Durairajan: Data collection and final approval of the version. Dinesh Durairajan: Data collection and final approval of the version. Divya Jilumudi, Harshavardhini Vasu, Vidya Viswanathan, Krishnaraj Karupasamy, Jerard Selvam, Selvavinayagam Thirumalaicheri Sivaprakasam, Arun Thamburaj, Vineeth S, Senthil Kumar, Prabhdeep Kaur: Conception of the work, critical review and final approval of the article.

